# Characterisation of ethnic differences in DNA methylation between UK resident South Asians and Europeans

**DOI:** 10.1101/2022.04.06.22273496

**Authors:** Hannah R. Elliott, Kimberley Burrows, Josine L. Min, Therese Tillin, Dan Mason, John Wright, Gillian Santorelli, George Davey Smith, Deborah A. Lawlor, Alun D. Hughes, Nishi Chaturvedi, Caroline L. Relton

**Author notes:** **Corresponding author:** Hannah R. Elliott. **Author email addresses:** Kimberley Burrows, Josine L. Min, Therese Tillin, Dan Mason John Wright, Gillian Santorelli, George Davey Smith, Deborah A. Lawlor, Alun D. Hughes, Nish Chaturvedi, Caroline L. Relton.

## Abstract

Ethnic differences in non-communicable disease risk have been described between individuals of South Asian and European ethnicity that are only partially explained by genetics and other known risk factors. DNA methylation is one underexplored mechanism that may explain differences in disease risk. Currently there is little knowledge of how DNA methylation varies between South Asian and European ethnicities.

This study characterised differences in blood DNA methylation between individuals of self-reported European and South Asian ethnicity from two UK-based cohorts; Southall and Brent Revisited (SABRE) and Born in Bradford (BiB). DNA methylation differences between ethnicities were widespread throughout the genome (n=16,433 CpG sites, 3.4% sites tested). Specifically, 76% of associations were attributable to ethnic differences in cell composition with fewer effects attributable to smoking and genetic variation. Ethnicity associated CpG sites were enriched for EWAS Catalog phenotypes including metabolites. This work highlights the need to consider ethnic diversity in epigenetic research.

## INTRODUCTION

There are well described ethnic differences in non-communicable diseases between migrants of South Asian genetic ancestry and their European ancestry counterparts^1^. The most striking difference is the greater risk of cardiometabolic disease in South Asians compared to Europeans (reviewed^2^). In contrast, all-cancer morbidity and mortality risks are lower in South Asian migrant groups compared to Europeans^3^. Ethnic differences in non-communicable diseases are only partially influenced by known genetic or environmental factors^4-6^. DNA methylation is one of the underexplored epigenetic mechanisms that may explain differences in disease risk.

In recent years, Epigenome Wide Association Studies (EWASs) have associated DNA methylation with diverse health-related outcomes such as cancer^7-10^, diabetes^11-13^, rheumatoid arthritis^14^, psychosis and schizophrenia^15^, blood pressure^16,17^, circulating metabolic measures^18^ and BMI^19-22^. DNA methylation has also proved a useful biomarker of lifestyle exposures, for example smoking^23-26^ and alcohol intake ^27,28^. Furthermore, DNA methylation may also be a predictor of future disease, for example myocardial infarction and coronary heart disease^29^ or all-cause mortality and time to death^30,31^. Ethnic differences have been explored in the context of these EWAS. For example, a meta-EWAS of 1,250 incident type 2 diabetes cases and 1,950 controls drawn from five European cohorts identified 76 CpG sites associated with incident type 2 diabetes^32^. Of these findings, 64 (84.2%) were directionally consistent in an independent cohort of Indian Asians. Evidence therefore suggests that many but not all DNA methylation associations with exposures or outcomes are consistent across populations indicating a possible role for DNA methylation as a mediator of excess disease risk between population groups.

Studies have begun to identify ethnic differences in DNA methylation patterns across the genome^33-35^. These studies highlight that ethnic differences in DNA methylation are likely to be frequent in the genome and Galanter et al.^33^ suggested they arise as a consequence of both genetic and environmental factors. However, to our knowledge there has been no large scale genome-wide comparison of DNA methylation in South Asian and European ethnic groups.

The aim of this study was therefore to identify and characterise DNA methylation differences between healthy adult individuals of self-reported South Asian and European ethnicity resident in the United Kingdom. We did this by first identifying cross-sectional differences in genome-wide DNA methylation using Illumina BeadChip arrays and then assessing stability of signals over time. We measured ethnic differences in DNA methylation derived estimates of cell composition and the methylation derived neutrophil to lymphocyte ratio (mdNLR), an index of systemic inflammation. We report the relative contribution of genetic and environmental sources of variance in ethnicity-associated DNA methylation and explore sources of environmental variation. We also assess whether ethnicity-associated CpG sites are functionally implicated in any biological pathways (using publicly available databases) or disease endpoints or risk factors captured by the EWAS Catalog^36^.

Analyses were conducted in the Southall And Brent Revisited (SABRE) cohort^2^ and replicated in the Born in Bradford (BiB) cohort^37,38^. These cohorts both include individuals of south Asian and Europeans in the UK but they are distinct in that they recruited from different geographical areas of the UK with South Asians in SABRE predominantly of Indian origin and South Asians in BiB predominantly of Pakistani origin. SABRE and BiB also differ in age, sex and other exposures, for example smoking behaviours. Inclusion of these two distinct cohorts is a strength of the study design, allowing identification of ethnicity associated DNA methylation sites that are likely to be generalisable.

## RESULTS

### Cohort Characteristics

A comparison of cohort and subgroup characteristics is shown in Table 1. All SABRE participants were male and recruited in middle age^2^. BiB participants were female, with DNA methylation measured on samples collected in pregnancy (24-28 weeks of gestation)^37,38^. Distributions of age, BMI and smoking differed between the two studies and for some measures, between the two ethnic groups (see Table 1).

**Table 1.** Cohort Characteristics

|  | SABRE Baseline |  |  | SABRE Follow-up |  |  | Born in Bradford |  |  | Cross cohort |  |  |
| --- | --- | --- | --- | --- | --- | --- | --- | --- | --- | --- | --- | --- |
| variable | European Mean (SD) or n (%) | South Asian Mean (SD) or n (%), | P-value<sup>a</sup> | European Mean (SD) or n (%) | South Asian Mean (SD) or n (%), | P-value<sup>a</sup> | White British Mean (SD) or n (%) | Asian Pakistani Mean (SD) or n (%), | P-value<sup>a</sup> | SABRE Baseline | BiB | P-value<sup>b</sup> |
| **N (%)** | 400 (50.0) | 400 (50.0) |  | 66 (47.5) | 73 (52.5) |  | 444 (48.5) | 472 (51.5) |  | 800 (46.6) | 916 (53.4) |  |
| **Males (%)** | 400 (100) | 400 (100) |  | 66 (100) | 73 (100) |  | 0 (0) | 0 (0) |  | 800 (100) | 0 (0) |  |
| **Age (years)** | 52.3 (7.1) | 51.0 (7.1) | 0.084 | 68.6 (5.3) | 69.0 (5.8) | 0.634 | 26.8 (6.1) | 28 (5.3) | 0.00205 | 51.9 (7.2) | 27 (5.7) | 0 |
| **BMI (kg/m<sup>2</sup>)** | 26.1 (3.7) | 26.0 (3.6) | 0.441 | 26.7 (3.1) | 26 (4.0) | 0.318 | 27.1 (6.3) | 26 (5.2) | 0.00269 | 26 (3.6) | 26 (5.8) | 0.0542 |
| **Bcell (%)** | 7.6 (2.09) | 9.1 (2.69) | 9.58E-20 | 8.3 (2.78) | 10.0(6.84) | 0.0424 | 4.75 (1.2) | 5.7 (1.5) | 1.62E-27 | 8.35 (2.5) | 5.3 (1.4) | 2.03E-151 |
| **CD4T (%)** | 14.5 (4.84) | 14.0 (4.32) | 0.00549 | 16.9 (5.74) | 15.0 (6.38) | 0.102 | 11.8 (2.6) | 12 (3.0) | 0.00104 | 14 (4.6) | 12 (2.8) | 1.50E-23 |
| **CD8T (%)** | 1.2 (2.23) | 2.1 (3.04) | 1.53E-06 | 0.5 (1.5) | 1.6 (2.74) | 0.00651 | 0.126 (0.7) | 0.36 (1.1) | 0.000122 | 1.66 (2.7) | 0.25 (0.9) | 1.24E-40 |
| **Eos (%)** | 0.06(0.368) | 0.26 (1.15) | 0.000598 | 0.2 (0.722) | 0.7 (1.78) | 0.0209 | 0.0185 (0.2) | 0.19 (0.8) | 1.57E-05 | 0.159 (0.9) | 0.11 (0.6) | 0.154 |
| **Mono (%)** | 10.3 (1.94) | 9.9 (1.69) | 0.00166 | 10.7 (2.04) | 9.5 (1.67) | 0.000175 | 11.2 (1.6) | 12 (2.01) | 0.00113 | 10.1 (1.8) | 11 (1.8) | 8.13E-45 |
| **Neu (%)** | 56.2 (8.52) | 53 (8.47) | 3.35E-06 | 53.9 (10.6) | 52 (13.2) | 0.308 | 68.7 (5.0) | 65 (6.97) | 1.77E-20 | 54.8 (8.6) | 67 (6.4) | 1.74E-171 |
| **NK (%)** | 15.8 (4.41) | 18 (4.65) | 8.97E-09 | 16 (4.7) | 18 (4.8) | 0.0227 | 6.55 (2.4) | 8.4 (2.74) | 8.51E-26 | 16.7 (4.6) | 7.5 (2.7) | 1.78E-295 |
| **ever smoking (%)** | 291 (72.8) | 127 (31.8) | 1.60E-30 | 45 (68.2) | 22 (31.1) | 1.60E-05 | 260 (58.6) | 35 (7.4) | 6.40E-61 | 418 (52.3) | 295 (32.2) | 1.80E-17 |

<sup>a</sup>t-test or $\chi^2$ for ethnic differences; <sup>b</sup>t-test or $\chi^2$ comparing SABRE baseline and Born in Bradford

### Concordance of self-reported ethnicity with genetic ancestry

Principal components analysis (PCA) of SABRE, BiB and HapMap3 populations was used to assign individuals to groups with similar genetic ancestry (Supplementary Figure 1).

Self-reported Europeans from SABRE and BiB cluster closely with Europeans from HapMap3 (Supplementary Figure 1). Correspondingly, self-reported South Asians from SABRE and BiB cluster with or near HapMap3 Gujarati Indians recruited from Houston, USA. PCA analysis on the self-reported South Asian and European individuals in SABRE and BiB show separate clustering by ethnicity. The genetic variation as estimated by genetic PCs appears higher in the South Asian group, where ethnicity subgroups can be separated on both the PC1 and PC2 axis. In SABRE, those indicating their ethnicity subgrouping as “Punjabi-” or “Gujarati” cluster more closely together than other subgroups.

### Concordance of self-reported ethnicity with DNA methylation components

Principal components (PCs) were derived from the SABRE and BiB methylation matrices. When utilising the 1,000 or 10,000 probes with greatest variance, PC1 differentiated between South Asian and European individuals in both SABRE and BiB. (Supplementary Figure 2) There was no differential clustering by ethnic group when using all independent probes from the array (n_SABRE_=21,023, n_BiB_=19,438).

### Identification of ethnic differences in DNA methylation

#### Ethnicity EWAS

In univariable analysis, differential methylation between SABRE individuals of self-reported South Asian or European ethnicity was identified at 32,435 CpG sites at p ≤1.03 × 10^−7^ (6.7% of the 484,781 sites assessed, Supplementary Table). Effect sizes ranged from 0.08% to 25.8% difference in DNA methylation between ethnic groups. CpG sites associated with ethnicity were distributed throughout the genome (Figure 1A and B), with hypermethylation amongst individuals of South Asian ethnicity being predominant (67% of CpG sites identified) when compared to Europeans (Figure 1A and C).

**Figure 1.**
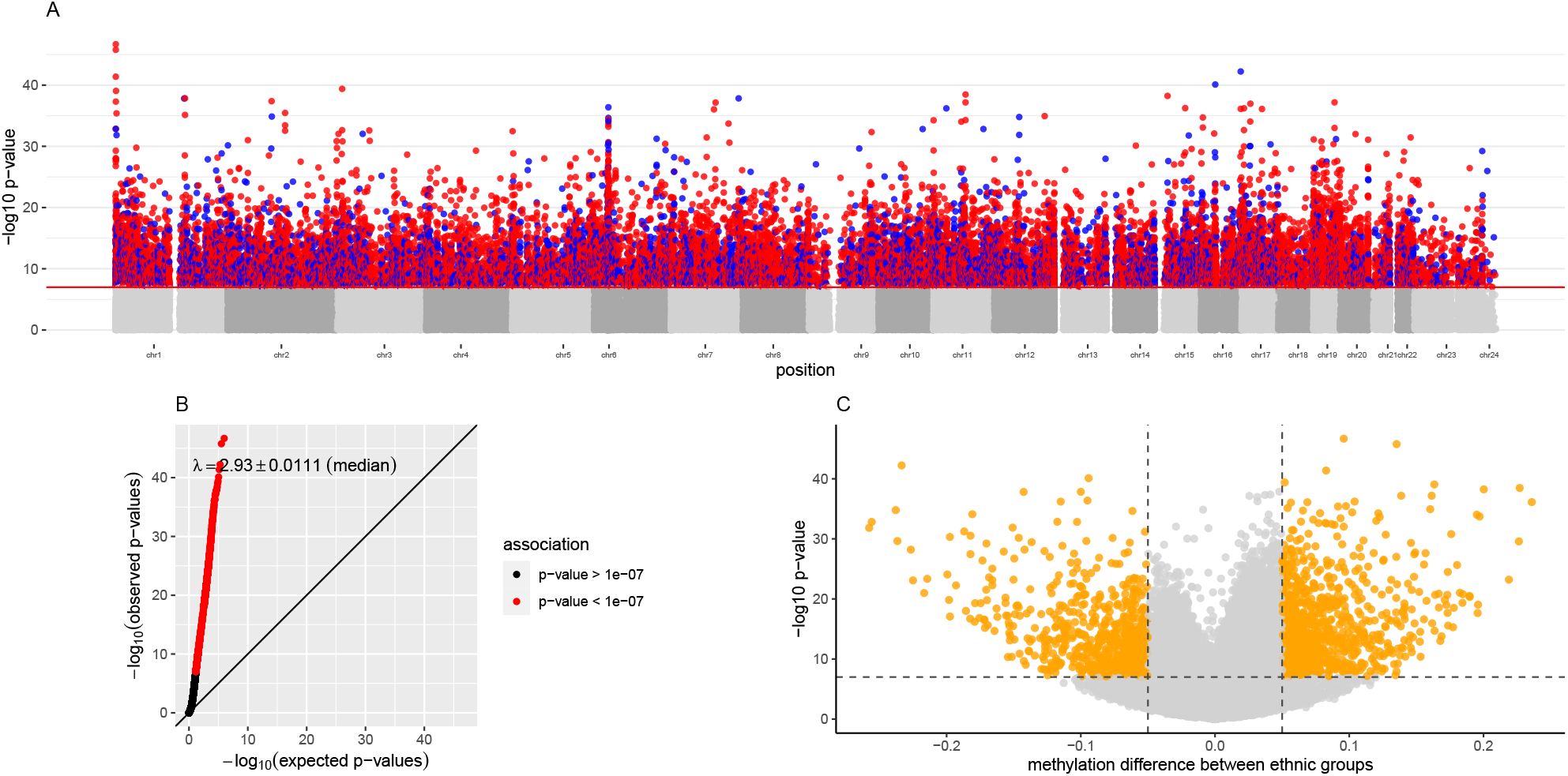
EWAS plots showing association between methylation and ethnic group in the SABRE cohort A: Manhattan plot showing association between ethnicity and DNA methylation. CpG sites with corresponding P-values at ≤1.03⍰×⍰10-7 are colour coded to show the direction of effect: red CpG sites are hypermethylated in South Asian individuals while blue CpG sites are hypomethylated relative to the European group. B: Q-Q plot showing observed vs expected p-values from the EWAS analysis. The red line denotes equality. λ= 2.93. C: Volcano plot showing p-value vs effect size for each of the tested CpG on the array. Orange highlighted CpGs are those with p<1.03⍰× ⍰10-7 and with effect sizes of >5%.

In BiB, effect estimates correlated with those reported in SABRE (R^2^=0.88, see Figure 2A). When considering P-values, 13,803/30,095 (46%) of the individual CpG sites were replicated as defined by maintenance of p-values below the epigenome-wide threshold (Supplementary Table).

**Figure 2.**
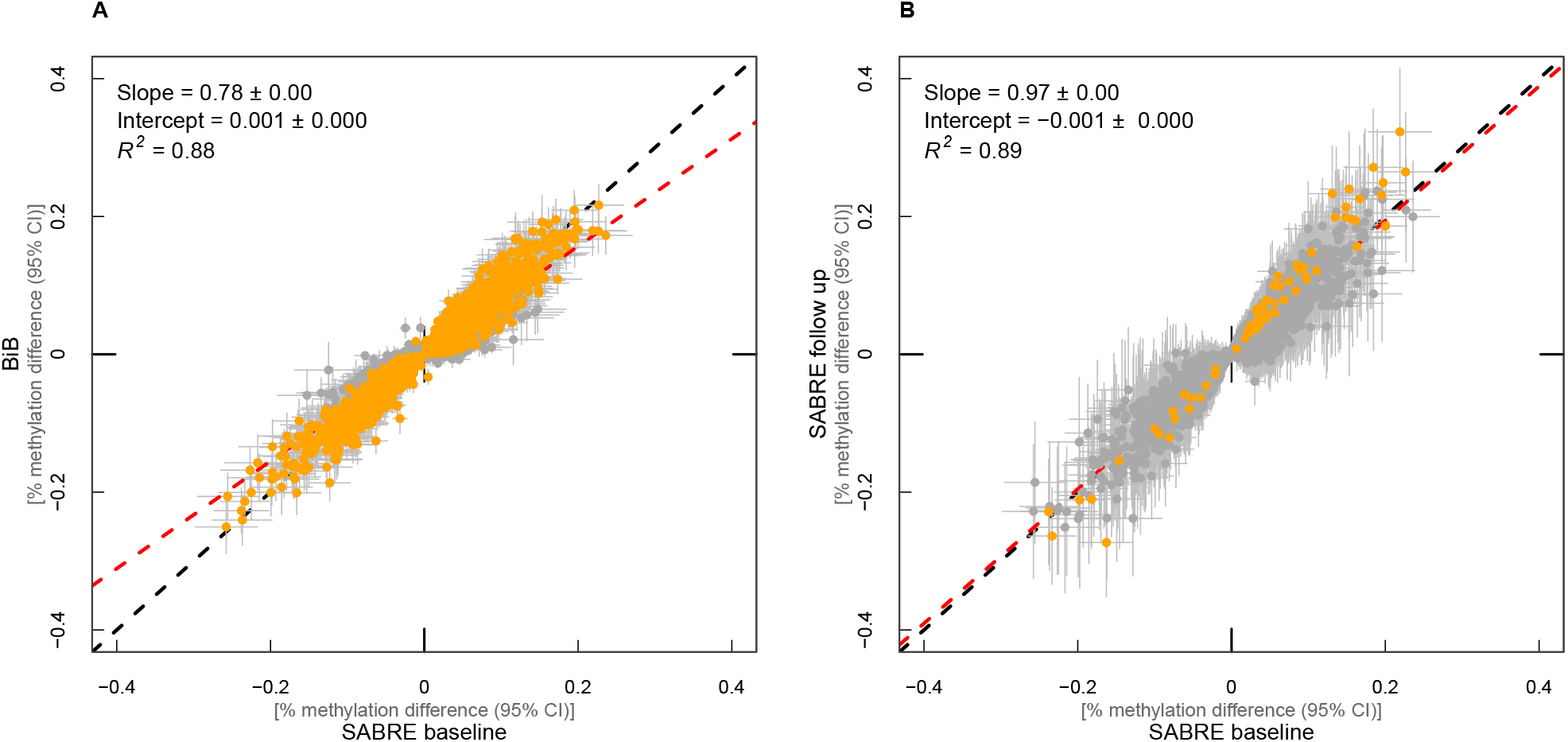
Correlation of ethnicity EWAS effect estimates between A) SABRE baseline and BiB individuals B) SABRE baseline and follow up timepoints Each CpG is represented by a point on the graph with 95% confidence intervals for effect estimates. Red dashed line: linear regression between datasets. Black dashed line: line of equality. Orange highlighted estimates: p ≤1.03 × 10^−7^ in BiB (A) or SABRE follow up timepoint (B) EWAS. A: N= 30,081 CpG sites (those associated with ethnicity from SABRE EWAS with data available in BiB. B: N=15,131 CpG sites associated with self-reported ethnicity at SABRE baseline and replicated in BiB.

We observed inflated λ values in both SABRE (λ=2.93) and BiB (λ=2.22) EWAS. λ values were recalculated described previously^39^ (see methods). λ was substantially reduced in both SABRE (λ=1.53) and BiB (λ=1.02), indicating inflation is likely to stem from abundance of true biological signal rather than substantial residual confounding.

#### Differentially Methylated Regions (DMRs)

DMR analysis was applied to the EWAS results to identify wider regions of DNA methylation that were associated with self-reported ethnicity. We identified 10,675 DMRs containing 2 or more CpGs that differed between the two ethnic groups at adjusted p<0.05 in SABRE (Supplementary Table 2 DMR results). Using the same analysis parameters, 5,371 (50%) of DMRs identified in SABRE had at least 1 overlapping CpG with DMRs identified in BiB (Supplementary Table 2 and Supplementary Table). The modal number of CpGs amongst replicated DMRs was 2 (n=1,870), and 73% of replicated DMRs contained fewer than 5 CpGs. The largest region identified in the DMR analysis was 1.1kb in length and contained 31 CpG sites. This region on chromosome 11 spans the upstream and exon 1 region of the imprinted *KCNQ1DN* locus and appears to contain CpG islands, DNase I hypersensitivity clusters, and transcription factor binding sites^40,41^.

All further analyses were restricted to a pool of 16,344 unique CpG sites: 13,803 replicated CpG sites identified in the univariable EWAS of ethnicity in SABRE and BiB and 2,541 CpGs representing replicated DMRs (the CpG with the lowest EWAS p-value selected from each DMR, Supplementary Table).

#### Ethnic differences in DNA methylation are stable over time

Participants in the SABRE study attended a follow-up clinic approximately 20 years after baseline recruitment (n=139), allowing an assessment of the temporal stability of ethnicity associated DNA methylation variation. Characteristics of individuals from the SABRE follow-up timepoint are shown in Table 1. Ethnicity EWAS effect estimates were compared between SABRE timepoints at replicated CpG sites (n=16,344 CpGs). Effect estimates correlated with those reported at SABRE baseline (R^2^=0.89, Figure 2B). Only 1685 (10.3%) CpG sites had effect estimates that changed by >2-fold between time points.

### Analysis of ethnic differences in methylation-derived phenotypes

DNA methylation data were used to generate an index of biological aging (using a variety of epigenetic clocks^42^), estimates of white blood cell counts^43^ and an index of systemic inflammation^44^. Subsequent analyses assessed whether these derived phenotypes differed between ethnic groups.

#### Ethnic differences in epigenetic age acceleration

Epigenetic age was computed using five clock algorithms (see Methods). SABRE South Asians had higher age acceleration estimates amongst all clock measures (except the Hannum clock) compared to SABRE Europeans in unadjusted tests (Supplementary Table 3). For both Extrinsic and Intrinsic Epigenetic Age Accelerations (EEAA and IEAA), these differences were robust to adjustment for BMI and self-reported smoking in PhenoAge (EEAA: beta=1.63, p=7.62×10^−5^; IEAA: beta=1.59, p=6.39×10^−5^) and SkinBlood estimators (EEAA: beta=0.75, p=1.50×10^−3^; IEAA: beta=0.75, p=9.78×10^−4^). Evidence for an association between age acceleration and ethnicity in BiB was weak (Supplementary Table 3).

#### Ethnic differences in cell composition

Using a linear model where estimated cell proportion was the outcome, ethnicity was the predictor and age was included as a covariate (Supplementary Table 4, model 1) we show that SABRE and BiB self-reported South Asian individuals had elevated estimated proportions of B cells, CD8+ T cells, Eosinophils and Natural Killer cells and reduced estimated proportions of Neutrophils compared to self-reported European individuals from the same cohort (Table 1). These relationships were unchanged when including smoking status as an additional covariate (Supplementary Table 4, model 2). Differences in the proportion of cell subgroups therefore exist between ethnic groups but these differences are not substantially explained by age or smoking status.

#### Ethnic differences in systemic inflammation

Methylation derived neutrophil to lymphocyte ratio (mdNLR), an index of systemic inflammation, was 13.6% lower in SABRE South Asians compared to Europeans (median (IQR): South Asians= 1.25 (0.70), Europeans=1.42 (0.73); p=2.47 ×10^−7^). This relationship was unchanged by adjustment for age, smoking and BMI. Similar findings were observed in BiB, with mdNLR 18.1% lower in South Asians compared to Europeans (median (IQR): South Asians=2.48 (1.04), Europeans=3.03 (1.14); p=1.59×10^−20^) in univariable analyses, and unchanged with adjustment for age, smoking or BMI.

### Determinants of ethnic differences in DNA methylation

To explore the potential factors mediating the relationship between self-reported ethnicity and DNA methylation we undertook a series of further analyses in SABRE.

#### The contribution of ethnic differences in smoking, age and cell composition to observed ethnic differences in DNA methylation

There were marked differences in self-reported smoking behaviour between SABRE ethnic groups and European participants were slightly younger than South Asian participants (see Table 1). Age and smoking status have both been shown to be associated with DNA methylation in previous studies^45,46^. Adjusting the univariable ethnicity EWAS model for age did not alter effect estimates (R^2^=1.0, Supplementary Table 1). Effect estimates when comparing the univariable ethnicity EWAS model with a model adjusted for age and self-reported smoking status were also highly correlated (R^2^ =0.98, Supplementary Table 1). However, the p-values of 2,995/16,344 (18%) of CpG sites were attenuated (p >1.03 × 10^−7^) following adjustment for age and smoking status; 9/16,344 (0.05%) of CpG sites had effect estimates that changed by >2-fold between models (Supplementary Table). Ethnicity-associated CpGs were compared to results from a large meta-analysis of smoking in adults^45^. We observed an increased proportion of smoking-associated CpGs amongst the ethnicity associated CpG sites in SABRE compared to expected (chi-square test: 23.5% vs 7.4%, p< 2.2e^-16^). We therefore conclude that smoking differences are likely to contribute to the observed ethnic differences in DNA methylation in up to 18% of CpG sites tested. Smoking associated CpGs^45^ are annotated in Supplementary Table.

Cell composition estimates were included as additional covariates in the univariable ethnicity EWAS model. In this analysis, effect estimates were correlated (R^2^=0.9) (Supplementary Figure 3). The p-values of 12,422/16,344 (76%) of CpG sites were attenuated following adjustment for cell subtypes (p >1.03 × 10^−7^) and 4,396/16,344 (26.9%) of CpG sites had effect estimates that changed by >2-fold between models (Supplementary Table, Supplementary Figure 3). Additional adjustment for age and smoking status had minimal effect: p-values of 12,580/16,344 (77%) of CpG sites were attenuated following adjustment for age, smoking status and cell subtypes combined (p >1.03 × 10^−7^) and 3,198/16,344 (19.6%) of CpG sites had effect estimates that changed by >2-fold between models (Supplementary Table). Differences in cell composition therefore appear to be an important driver of ethnicity associated methylation patterns and these differences are not strongly driven by differences in age or smoking status between ethnic groups.

#### Genetic contribution to ethnic differences in DNA methylation

##### Assessment of genetic principal components

Genetic principal components generated by PCA (n=20, see methods) were included as additional covariates in the univariable ethnicity EWAS model. None of the 16,344 ethnicity-associated CpG sites had p values at p≤1.03⍰×⍰10^−7^ following adjustment for genetic principal components.

##### Assessment of mQTLs

Using the Genetics of DNA Methylation Consortium (GoDMC) data^47^, 16,720 mQTLs were identified (containing 15,566 unique SNPs and 10,171 unique CpGs)^47^. 62% of ethnicity associated CpGs had at least one mQTL (10,171 of 16,344 CpGs queried, CpGs annotated in Supplementary Table). In GoDMC, 45% of tested CpGs had an mQTL, ethnicity associated CpGs in this study therefore appear to be enriched for mQTLs (OR=1.00, 95%CI=1.93-2.06, p=0). A total of 8,686 SNPs were available in the SABRE cohort which allowed us to further explore 9,382 mQTLs across 8,318 unique CpGs.

We tested each available SNP to identify differences in allele frequency between SABRE ethnic groups. We estimate that 2,908 (33.5%) of the SNPs tested differ in allele frequency between populations (adjusted p<0.05, n_tests_=8,686) (Supplementary Table 5, annotated in Supplementary Table). To investigate further, we included the respective mQTL genotype as a covariate in the univariable ethnicity EWAS model for each CpG with an mQTL (Supplementary Table 5). Variance explained (measured by R^2^) by the models increased by a mean of 2.7% (SD= 5.3%) in CpGs with differences in allele frequency between populations and a mean of 1.7% (SD= 3.2%) in CpGs showing no differences in allele frequency between populations.

We therefore conclude that ethnic differences in DNA methylation may be partially attributable to differences in allele frequency between ethnicities but overall the contribution of these genetic effects are small. Amongst CpGs with mQTLs that varied in allele frequency between ethnic groups, adjusting for the mQTL attenuated the EWAS signal above the EWAS p-value threshold in only 763/2,908 (26.2%) of unique CpG sites (Supplementary Table 5).

We also postulated that DNA methylation differences between ethnic groups may be driven by differences in mQTL effect between ethnicities. We therefore tested each available SNP to identify interactions between mQTL and ethnic group on methylation using results from the GEM interaction model^48^. GxE models were run where methylation was the outcome variable, genotype x ethnicity was the predictor and age and smoking were included as covariates. We identified 67 interactions (adjusted p<0.05, n_tests_=9,382) between ethnicity and mQTLs indicating that a small number of CpG associations with ethnicity may be attributable to ethnic specific genetic effects (Supplementary Table, Supplementary Table 6). None of the 61 unique interaction mQTLs overlap with a multi-ethnic GWAS of cell composition^49^. We also compared the 66 unique CpG sites showing evidence of interaction between ethnicity and mQTLs with the univariable EWAS and the EWAS model which included cell composition as a covariate. The p-values of 15/66 (23%) of CpG sites were attenuated (p >1.03 × 10^−7^) following adjustment for cell composition but none of the effect estimates changed by >2-fold between models (Supplementary Table). This indicates that interactions between ethnicity and mQTLs are not related to differences in cell composition.

### Functional annotation of ethnicity associated CpG sites

We initially identified a pool of 16,344 unique CpG sites that were associated with ethnic group in SABRE and replicated these in BiB. To characterise them in terms of biology and disease relevance we undertook a series of further analyses.

#### Ethnicity associated CpGs are depleted for CpGs in 3’UTRs and intronic regions

Ethnicity associated CpGs were depleted for CpGs in 3’UTRs (OR= 0.71, 95% CI= 0.64-0.78, p=1.19 × 10^−12^) and depleted for CpGs in intronic regions (OR=0.90, 95% CI= 0.87-0.93, p=2.32 × 10^−10^) based on ANNOVAR annotation compared to all CpGs on the 450k array (Supplementary Table 7 ANNOVAR enrichment).

#### Ethnicity associated CpGs are enriched in multiple phenotype groups based on EWAS Catalog data

Results from the EWAS Catalog^36^ were grouped into related phenotypes^50^ and tested for enrichment. The 16,344 ethnicity associated CpGs are strongly enriched for CpG sites previously reported to be associated with metabolites (OR= 4.12, 95%CI= 3.64-4.67, p= 3.14 × 10^−129^), smoking (OR= 2.20, 95%CI= 2.06-2.35, p= 1.13 × 10^−130^), alcohol (OR= 2.10, 95%CI= 1.80-2.43, p= 1.47 ×10^−23^), cancer (OR=1.83, 95%CI= 1.77-1.90, p= 4.11 × 10^−251^) and perinatal phenotypes (OR= 1.89, 95%CI= 1.58-2.26, p= 1.00 ×10^−12^) and strongly depleted for autoimmune (OR=0.34, 95%CI=0.32-0.36, p=1.31 × 10^−278^) and infection phenotypes (OR=0.52, 95%CI= 0.48-0.57, p=1.02 × 10^−52^) compared to all entries in the EWAS Catalog. CpG sites are also enriched for CpG sites associated with ancestry (OR=1.50, 95%CI= 1.24-1.82, p=1.80 × 10^−5^) (Figure 3).

**Figure 3.**
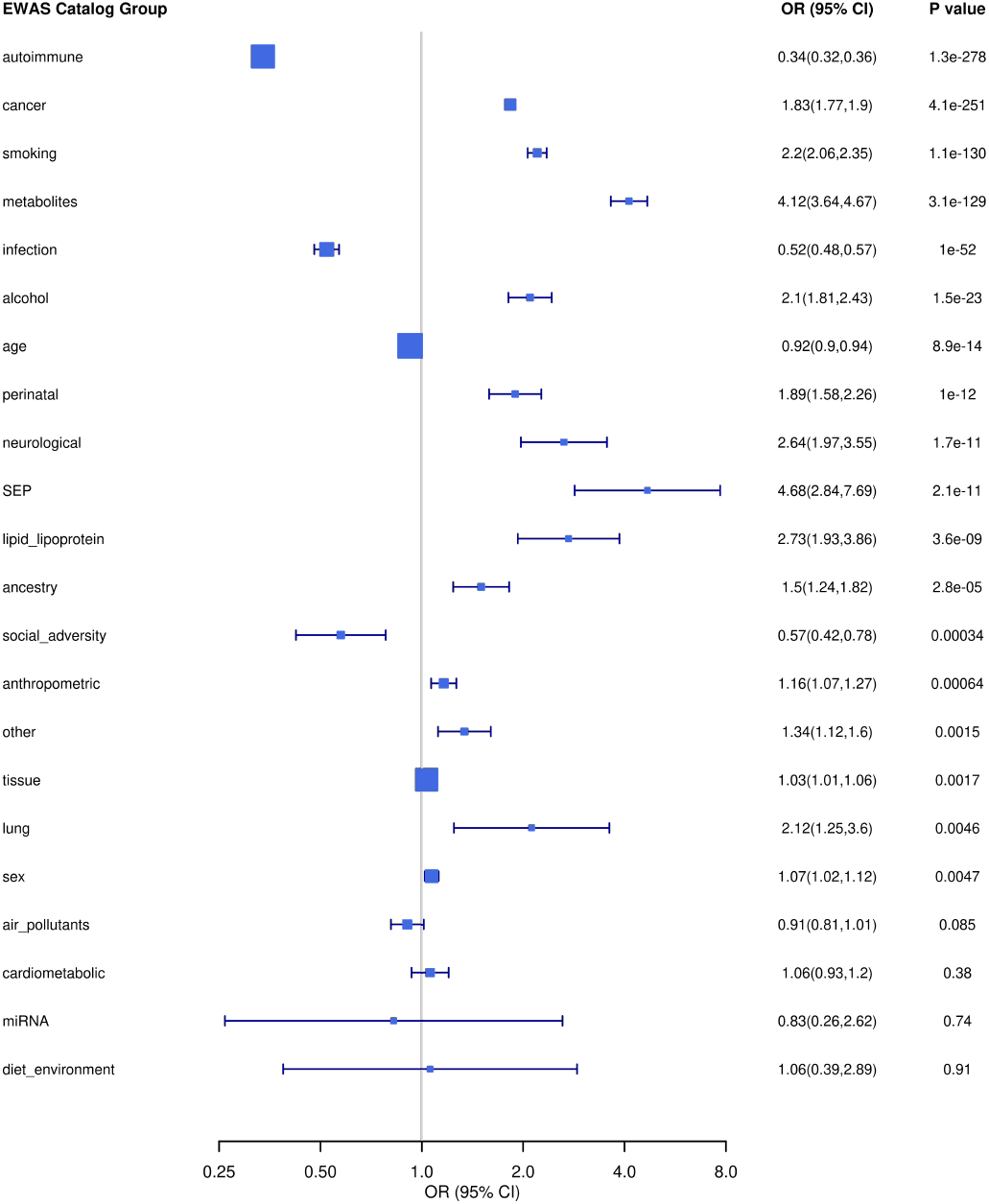
Enrichment of EWAS Catalog phenotypes amongst ethnicity associated CpG sites Entries from the EWAS Catalog were reduced into categories of related phenotypes^50^. Group “other” contained CpGs not assigned categories and represented 0.24% of all unique CpGs across categories. SEP: Socio-economic position.

#### Ethnicity associated CpG sites are highly enriched for cis-eQTMs

1,372 ethnicity associated CpGs were expression quantitative trait methylation (eQTMs) based on BIOS data^51^ and ethnicity associated CpGs were highly enriched for eQTMs compared to the overall BIOS dataset (OR=2.72, 95% CI= 2.57, 2.88, p= 6.78 × 10^−271^). eQTMs are annotated in Supplementary Table.

#### Ethnicity associated CpG sites are enriched for several GO terms

There were 427 GO terms enriched at FDR <0.05 amongst hyper-methylated CpGs. There was enrichment for Biological Process terms including several related to morphogenesis and development. Others were related to synaptic signalling. There was also enrichment for Cellular Component terms “integral/intrinsic component of plasma membrane”, “plasma membrane”, “cell periphery” (Supplementary Table 8). There were 50 GO terms enriched at FDR <0.05 amongst hypo-methylated CpGs. Enrichment of Biological Process terms were predominantly terms related to immune response. Enrichment of Cellular Component terms were related to “cell surface” and “plasma membrane” (Supplementary Table 8).

We repeated the GO ontology enrichment analysis, this time restricting to CpGs that were identified as eQTMs based on BIOS data (Supplementary Table 8). In this analysis, hypermethylated CpGs were enriched for 18 GO terms, almost all were Molecular Function terms. The most strongly enriched terms were “transcription regulatory region sequence-specific DNA binding”, “RNA polymerase II transcription regulatory region sequence-specific DNA binding”, “RNA polymerase II cis-regulatory region sequence-specific DNA binding” and “regulatory region nucleic acid binding”. Amongst hypo-methylated CpGs, we detected enrichment of 54 GO terms predominantly of Biological Process terms relating to immune response (Supplementary Table 8 GO term enrichment).

#### Regulatory pathways overlap with the genomic position of ethnicity associated CpGs

Using LOLA, we identified regulatory elements overlapping with the genomic position of hyper- and hypomethylated sets associated with ethnicity (Supplementary Table 9). Overlaps in the hypermethylated set includes sites annotated to CpG islands, DNAse hypersensitivity sites across multiple tissues including stem, brain, liver and hematopoietic cells and transcription factor binding sites (most strongly EZH2, a component of the polycomb repressive complex 2 (PRC2) which functions to methylate Lys-27 on histone 3). The hypermethylated set also includes overlaps with histone modifications with the strongest evidence for overlaps with H3K4me1 and H3K27me3 which indicate active enhancer regions^52^. This is also supported by overlaps with enhancer segments across multiple cell types.

In the hypomethylated set, the overlaps were fewer than in the hypermethylated set (n=74 vs n=299) Overlaps in the hypomethylated set include those with repressed segments across multiple cell types and UCSC repeat regions. DNAse hypersensitivity site overlaps in the hypomethylated set were observed predominantly in hematopoietic cells. Transcription factor binding site overlaps include c-Fos and C-Jun which combine to form activator protein-1, a transcription factor which coordinates transcription in response to cytokines and infection by viruses and bacteria^53^. In contrast to the hypermethylated set, there were no overlaps with histone modifications in the hypomethylated set. (see Supplementary Table 9).

## DISCUSSION

We observed genome-wide differences in DNA methylation between South Asian and European individuals at 16,344 CpG sites and regions with effect sizes that were comparable in magnitude of effect to those seen in EWAS of smoking^23,25,45^. These differences were replicated between two distinct cohorts; one of men in older adulthood and one of women in pregnancy. By using DNA samples given at follow up clinics in SABRE, we showed temporal stability of ethnic differences in DNA methylation over a c.20 year period. We also observed lower levels of systemic inflammation (measured by mdNLR) in South Asians compared to Europeans in both cohorts. Ethnic differences in epigenetic age acceleration were found for some of the estimators tested (namely PhenoAge and SkinBlood), but only in SABRE. Functional exploration of the ethnic differences observed indicated that ethnicity associated CpG sites are depleted in in 3’UTRs and intronic regions and highly enriched for eQTMs. By comparing ethnicity associated CpG sites with EWAS Catalog entries, we identified that ethnicity associated CpGs are enriched amongst multiple groups of traits including metabolites and autoimmune phenotypes, that had previously been reported to be associated with variation in DNA methylation. On further investigation within the SABRE cohort, we found that ethnic differences are predominantly explained by differences in cell composition, and to a lesser extent explained by smoking and genetic effects.

Initial exploration of genetic and DNA methylation variance using PCA demonstrated that individuals from each self-reported ethnic group were genetically and epigenetically distinct. Genetic variation estimated from genetic PCA is greater in South Asian compared to European individuals in SABRE and BiB. This is expected since South Asian individuals in these two cohorts have a recent migration history from a large geographical area into the United Kingdom. Methylation PCA analysis also separated self-reported ethnicities but this was less distinct than for genetic data.

Ethnicity associations in SABRE were temporally stable (effect estimates R^2^=0.9 over 20 years) although we acknowledge there may be a healthy survivor effect since around 25% of the cohort died between baseline and follow up clinics. In previous studies, CpG sites stable over time have been postulated to be driven by genetic effects^54^. However, our data suggest that genetic variation is not a strong determinant of ethnicity-associated differences in DNA methylation. We therefore suggest that the DNA methylation differences observed are likely to be predominantly environmentally determined and persist across the life course. For example, this hypothesis is consistent with reported observations of early life exposures leaving a lasting impact on the methylome^55^.

A number of studies have identified differences in epigenetic age acceleration between ancestries^56-58^, although none has previously studied South Asian individuals. Self-reported South Asian individuals had higher age acceleration amongst all epigenetic clock measures than those in the European ethnic group in SABRE. For Levine PhenoAge and Horvath SkinBlood estimators these differences were robust to adjustment for BMI and self-reported smoking. We did not replicate results in BiB but postulate this may be due to the younger age of BiB participants. It has been suggested that epigenetic clocks may contain ancestry specific CpGs in their models so further exploration of clock models is required before concrete inferences can be made with respect to the biological or clinical significance of our observations^59^.

An index of systemic inflammation derived from relative proportions of white blood cell types (mdNLR) was substantially lower in South Asians compared to Europeans in both SABRE and BiB and the association was not explained by age, smoking or BMI. Higher mdNLR has previously been associated with increased cancer^60-62^, rheumatoid arthritis^63^ and cardiovascular disease risk^64^. In previous studies, differences in mdNLR by ethnicity have been reported with European ancestry groups having the highest level of mdNLR^44,65^. Our results include South Asian ancestries for the first time and support Europeans having elevated mdNLR compared to other ancestries. The presence of differences between ethnicities in this cell count derived measure accords with observations in this study of cell counts being the major determinant of ethnic differences in DNA methylation, as discussed below.

Exploration of EWAS results identified that ethnic differences in cell composition were the major driver of ethnic differences in DNA methylation between South Asians and Europeans. We estimated that 76% of all CpG sites associated with ethnicity were attributable to differences in cell composition. This highlights the importance of accounting for cell composition in epigenetic studies, especially when there may be heterogeneity of genetic ancestries within a study.

We also estimated that 18% of the ethnic differences in DNA methylation observed between SABRE South Asians and Europeans were attributable to smoking. We note we used self-reported never vs ever smoking in analyses. Use of a more granular measure such as smoking pack years may capture more residual confounding but this finding underscores the need to adjust for common confounders in epigenetic studies.

We investigated the contribution of common genetic (SNP) effects to observed differences in DNA methylation between ethnic groups. In line with other studies^47^ we identify that mQTL effects are common but small in magnitude. In addition, we identified 67 interaction effects between mQTLs and ethnicity indicating there may be associations between methylation and ethnicity that are attributable to ethnic-specific genetic effects. These findings are of interest and require further exploration particularly given the mQTL reference panel consisted of European ancestry individuals. We may not have therefore captured SNP effects specific to individuals of South Asian ethnicity within SABRE.

In addition to exploring the potential determinants of DNA methylation differences between Europeans and South Asian ancestral groups, we sought to investigate whether there was any evidence to support the hypothesis that these ethnicity-variable CpG sites may be associated with biological functions (using GO, eQTM and LOLA) or disease phenotypes or risk factors (using the EWAS Catalog). We acknowledged earlier that European and South Asian ethnic groups show marked discordance in their risk for some non-communicable diseases, and we postulate that variation in DNA methylation may contribute to this. We show that ethnicity associated CpG sites are not randomly distributed throughout the genome and are more likely to be eQTMs than chance alone. Using LOLA, we identified overlaps between regulatory elements and CpG sites associated with ethnicity suggesting ethnicity associated CpG sites were enriched in active enhancer regions. These findings suggest that ethnicity associated CpG sites are enriched for functionally important sites.

Comparing ethnicity associated CpG sites with those in the EWAS Catalog^36^, we identified enrichment for CpG sites associated with ancestry. This gives further support to our EWAS results. We also observed enrichment amongst CpG sites associated with sex and smoking. Since our analyses are stratified by sex and adjusted for smoking this may suggest residual confounding or highlight that other studies reporting their associations in the EWAS Catalog may have not adjusted for these covariates. We observed strong enrichment amongst CpG sites associated with disease phenotypes, most prominently an enrichment of ethnicity associated CpG sites associated with metabolites and a depletion of ethnicity associated CpG sites amongst CpG sites associated with autoimmune diseases amongst others. This finding supports an avenue of further research to define ethnic specific risk. However, SABRE samples are restricted to healthy individuals (e.g. with no existing diabetes or cardiovascular disease) so this limits the conclusions we can make on phenotype enrichment compared to the EWAS Catalog data.

### Strengths and limitations

This is one of the largest studies to explore differences across the epigenome between South Asian and European populations and makes an important contribution to an emerging but still small literature on ethnic differences in epigenetic markers. We have also replicated epigenome wide ethnic differences in an independent cohort (BiB). That the replication sample (women only with DNA methylation measured on pregnancy samples) was very different to the discovery sample (men only in older age) has some advantage, in that where replication occurs this is likely to reflect robust ethnic differences. Additional strengths include the availability of genomic and other data in the two studies that enabled exploration of factors relating to ethnic differences in DNA methylation.

We acknowledge some key limitations. All reference data utilised is predominantly from datasets of European ancestry. For example, mQTLs from GoDMC were identified in individuals of European ancestry^47^. Therefore, we may not have identified mQTLs specific to individuals of South Asian ancestry which would mean that genetic effects may be underestimated in our study. EWAS studies in the EWAS Catalog are also predominantly conducted in European ancestry individuals. There may also be bias in methylation derived variables (cell composition and mdNLR) which rely on a European ancestry reference set^43,44^. The main EWAS was analysed in male participants and replication in females. There may therefore be sex-specific ethnic differences that we could not detect in this analysis. In addition, both SABRE and BiB samples are restricted to selected subsets of healthy individuals (see methods). This potentially introduces selection bias^66^ and limits the conclusions we can make on phenotype enrichment compared to the EWAS Catalog data. Finally, individuals in SABRE and BiB were recruited in the UK, with South Asian individuals representing migrant populations. This may limit generalisability to non-migrant South Asians vs Europeans.

This study has only made comparisons between two self-reported ethnic groups and models a limited number of phenotypes. Wider efforts are required to increase population diversity in epigenetic studies.

### Conclusions

This study aimed to define and characterise differences in DNA methylation between individuals of European and South Asian self-reported ethnicity. This is important research because there is little epigenetic data collected across global populations and very few cross-ancestry comparisons made. This study highlights widespread differences in DNA methylation throughout the genome between South Asians and Europeans. We estimate that 76% of ethnic differences observed in this study are attributable to differences in cell composition. Surprisingly little ethnicity-associated variation in DNA methylation was explained by underlying genetic differences between the groups. We also identified that South Asians have lower levels of systemic inflammation (using the methylation derived neutrophil to lymphocyte ratio) but in the older SABRE cohort we also observed higher age acceleration in South Asian individuals compared to Europeans. We used public databases to characterise ethnicity-associated CpG sites and show they are enriched for eQTMs and GO terms, depleted in 3’UTRs and intronic regions and have overlaps with regulatory markers. This study demonstrates that epigenetic differences between ethnicities are widespread. It highlights the need to consider cell composition, genetic variation and lifestyle confounders in population studies, particularly in studies with participants from different ancestries. This study also identifies that there are many CpG sites that are associated with methylation independently of these factors. Further exploration of these ethnicity-associated CpG sites could improve our understanding of disease aetiology and refine predictive models which rely on epigenetic data.

## METHODS

### Study populations

#### SABRE

Samples were derived from the extensively characterised population-based Southall And Brent REvisited (SABRE) cohort^2^. The SABRE cohort includes 1,711 first generation South Asian migrants and 1,762 European origin individuals aged between 40-69 living in west London, UK. Initial investigations were carried out between 1988 and 1991 with follow up clinics c.20 years later.

DNA was extracted from peripheral blood samples collected at baseline and follow up visits. A random sample of 800 men from the SABRE cohort at baseline clinic was selected for this study. Sampling was limited to individuals free from diabetes or coronary heart disease with good quality DNA available at baseline. Samples were stratified across two age groups (cut at the median of 52 years) and across two self-reported ethnic groups (South Asian and European). Samples were randomly selected to give equal numbers of samples in each stratum. Repeat DNA was collected at follow-up clinics where possible (n=139/800 individuals).

The study was approved by St Mary’s Hospital Research Ethics Committee (07/H0712/109) and all participants provided written informed consent.

#### Born in Bradford

Born in Bradford (BiB) is a multi-ethnic pregnancy and birth cohort that recruited pregnant women largely during an oral glucose tolerance test (24-28 weeks gestation) in Bradford, UK between 2007 and 2010^37^. The cohort has been described in detail elsewhere^37,38,67^. In total 12,453 women, who gave birth to 13,818 infants were recruited. DNA methylation was generated from a non-random subsample of 1000 BiB maternal DNA samples, that prioritised the two largest (self-reported) ethnic groups (White British and Pakistani) and included women who had a singleton pregnancy and where both mother and their child had genome-wide data. Samples were selected to give equal numbers of self-reported white British (n=444) and Pakistani (n=472) mother-offspring pairs.

Ethical approval for the data collection was granted by Bradford Research Ethics Committee (Ref 07/H1302/112).

### Methylation data

DNA from SABRE participants was analysed using HumanMethylation450 BeadChips (Illumina, San Diego, CA, USA). Approximately 500ng of DNA was bisulphite modified using EZ DNA methylation kits (Zymo Research, Orange, CA, USA). The manufacturer’s protocol was followed using the alternative incubation conditions recommended when using Illumina BeadChips. BeadChips were processed according to the manufacturer’s protocol and Illumina iScan software v3.3.28 was used to scan the arrays. Quality control and processing was conducted according to the pipeline described previously^68^ but with more stringent quality control parameters: The detection p value threshold was reduced to 0.1 for samples and probes, bead number threshold was reduced to 0.1 for samples and probes, sample genotype concordance was reduced to 0.8 and sex outliers <5 SD were removed. In total, 11 samples and 1644 probes were removed from the dataset. SABRE data were normalized using 15 control probe PCs derived from the technical probes informed by *meffil* scree plots^68^. During normalisation of raw data, batch effects (defined as 450k slide ID) were removed where slide was modelled as a random effect. Cell composition was estimated from methylation data using the Houseman method^43,69^. Statistical analyses were performed on β-values throughout. Samples with missing genetic data or admixture (Supplementary Figure 1) were also removed leaving 747 samples and 484,781 probes available for analysis.

In BiB, the data generation pipeline was analogous to that of SABRE, but the methylation array used was the HumanMethylationEPIC BeadChip (Illumina, San Diego, CA, USA). Following QC steps, 922 samples and 860,750 probes were available for analysis.

#### Methylation principal components

CpG sites were pruned by ranking on variance and removing CpGs with correlation R^2^>0.2. In each correlated pair the CpG with the highest variance was retained. This pruning resulted in a reduced dataset of 21,023 CpGs (SABRE) and 19,438 CpGs (BiB). Principal component analysis (PCA) then was performed to generate methylation principal components using *prcomp*.

### Genetic Data

DNA from 2980 SABRE participants were genotyped using the UCL druggable target array, comprising the Illumina Human 1679 Core Bead Chip (∼240K genome wide markers) and an additional custom set of 200K markers on genes encoding proteins involved in drug handling, drug action, and druggable targets. This was developed in collaboration with London School of Hygiene and Tropical Medicine and the European Bioinformatics Institute.

For both the SABRE South Asian and European sub-cohorts, individuals were excluded on the basis of incorrect sex assignment, high missingness (>5%), abnormal heterozygosity (het > mean (het) + 3*sd (het) or het < mean (het) - 3*sd (het)), cryptic relatedness (pihat>=0.9999 identified 20 cases of sample mislabelling which were excluded) and non-concordant ancestry (detected via Principal Components Analysis).

SABRE phasing was performed using Eagle and ethnicity-specific imputation was performed using Minimac3 against the HRC reference panel (http://www.haplotype-reference-consortium.org/)^70^.

SABRE genotypes were filtered to have a MAF>0.01, imputation info score>0.8 and HWE p<0.00001. After data cleaning and QC, 1,527 participants of European ethnicity and 1,210 participants of South Asian ethnicity had available 6,912,559, and 6,046,044 SNPs respectively.

For BiB participants, DNA was genotyped using either Infinium Human Core Exome-24 v1.1 arrays or Infinium global screen-24+v1.0 arrays (Illumina, San Diego, CA, USA). Samples were pre-processed using GenomeStudio 2011.1. Samples with Call Rate<0.95 were excluded. Poorly performing SNPs were identified based on Call Freq<0.97, Cluster Sep≤0.3, AB R Mean≤0.2, BB R Mean≤0.2, AA R Mean≤0.2, 10% GC Score≤0.2, MI Errors<2 and Rep Errors<2.

After quality control, a VCF containing 15,628 BiB participants and 459,340 genomic variants was submitted to the Sanger Imputation Service using the “UK10K + 1000 Genomes Phase 3” as a reference panel and “pre-phase with EAGLE2 and impute” as the pipeline. The 1000 Genomes Phase 3 panel was chosen as it contains samples originating from several global populations^71^. The resultant imputation dataset contains 15,628 participants and 87,558,135 variants.

#### Genetic Principal Components

For the SABRE cohort, we combined European and South Asian populations on intersecting imputed SNPs (n= 5,539,760) and availability of DNA methylation data. In total, 749 European and South Asian individuals were included in the genetic principal component analysis.

For the BiB cohort, there were 875 individuals after sub-setting for participants with available DNA methylation and genetic data.

The HapMap Phase 3 (HapMap3) reference dataset contains around 1.5 million SNPs genotyped in 1,397 individuals from a variety of populations. This dataset is available from the HapMap FTP site (ftp://ftp.ncbi.nlm.nih.gov/hapmap/). We used LiftOver to convert HapMap3 genomic positions from Build 36 to 37 (hg18 to hg19 chain file) and merged with the SABRE and BiB cohort independently on intersecting SNPs (N= 1,032,847 (SABRE) and N= 1,348,525 (BiB)) giving a final sample of 2,146 individuals for SABRE and HapMap3 combined, and 2,272 individuals for BiB and HapMap3 combined.

Principal component analysis (PCA) was performed to generate the first 20 genetic principal components (PCs) for i) the SABRE cohort and BiB cohorts (independently) and ii) each cohort merged with HapMap3 using Plink 1.90^72^. There were 1,032,847 SNPs intersecting between SABRE and HapMap3 and 1,348,525 SNPs intersecting between BiB and HapMap3. After pruning for independent SNPs (window size=50 SNPs, step size=5 SNPs, VIF=1.5, and excluding long range LD regions); 103,973 and 120,671 SNPs were taken forwards for PCA in the SABRE cohort and combined SABRE and HapMap3 respectively. For BiB, there were 212,061 and 252,916 SNPs taken forwards for PCA in the BiB cohort and combined BiB and HapMap3 respectively.

### Derived Phenotypes

Measures of Epigenetic age and age acceleration were generated using the *methylclock* R package^42^. The following estimators were utilised: Horvath, Hannum, Alfonso and Gonzalez, Horvath Skin and Blood, Levine PhenoAge^73-78^. Extrinsic Epigenetic Age accelerations (EEAAs) for each method were calculated as the residuals from a linear model where DNAmAge was the outcome and chronological age was the predictor. Intrinsic epigenetic age accelerations (IEAA) were calculated as the residuals from a linear model, where DNAmAge was used as the outcome and chronological age and cell counts were predictors^30^.

The methylation derived neutrophil to lymphocyte ratio (mdNLR) is an epigenetic estimate of neutrophil to lymphocyte ratio^44^. Using cell count estimates, the neutrophil count was divided by the lymphocyte cell count (calculated as the sum of B cells, CD4+ T cells, CD8+ T cells and natural killer cells) to provide the mdNLR.

### Statistical analysis

Baseline characteristics comparing South Asians and Europeans were conducted using t-tests for continuous and chi-squared tests for categorical variables.

EWAS were conducted utilising *meffil*, where methylation was the outcome and ethnicity the predictor. Covariates in each model are described in the results. Differentially methylated regions (DMRs) were identified by combining EWAS test statistics between consecutive CpG sites (maximum distance between sites=500 base pairs) using the R package *dmrff*^79^.

To replicate our findings in an alternative cohort, we conducted an EWAS and DMR analysis of self-reported ethnicity amongst individuals from the Born in Bradford cohort.

To investigate inflation of observed λ values we used a method described previously^39^ where lambdas were recalculated in SABRE using CpGs which had p>0.2 in the corresponding BiB EWAS and vice versa where lambdas were recalculated in BiB using CpGs which had p>0.2 in the corresponding SABRE EWAS.

For epigenetic age analyses, linear models were used to identify differences in age acceleration between SABRE and BiB ethnic groups where ethnicity was the predictor and age acceleration was the outcome. In adjusted models, BMI and smoking were included as additional covariates in the models as these are common disease risk factors and differed by ethnicity in SABRE and BiB.

For mdNLR analyses, linear models were used to identify differences in mdNLR between SABRE and BiB ethnic groups where ethnicity was the predictor and mdNLR was the outcome. In adjusted models, BMI, smoking and age were included as additional covariates in the models as they are possible mediators in the relationship between ethnicity and mdNLR.

The Genetics of DNA Methylation Consortium (GoDMC) database (http://mqtldb.godmc.org.uk/)^47^ was used to identify mQTLs of ethnicity associated CpG sites identified in SABRE. The mQTLs retrieved were restricted to those at p<1 × 10-8 (cis mQTLs) or p<1 × 10-14 (*trans mQTLs*) in accordance with the GoDMC meta-analysis protocol^47^. For SNP analyses, allele frequencies were compared between populations using Fisher’s tests. In regression models, SNPs were coded as additive effects.

Genetic associations with methylation and gene environment interactions were modelled using *GEM*, using self-reported ethnicity as the environmental component^48^.

Genomic region information was annotated through ANNOVAR^80^. CpG sites were annotated into functional categories including: downstream, exonic, exonic/splicing, intergenic, intronic, ncRNA_exonic, ncRNA_exonic/splicing, ncRNA_intronic, ncRNA_splicing, splicing, upstream upstream/downstream, UTR3, UTR5, UTR5/UTR3. Enrichment was assessed using chi-squared tests.

Ethnicity associated CpG sites were compared to those listed in EWAS Catalog (http://www.ewascatalog.org/) in June 2021. The EWAS Catalog contains EWAS studies analysing at least 100,000 CpG sites using a minimum sample size of 100 individuals. Catalog entries were reduced into related categories^50^ and enrichment assessed using Wald odds ratio and chi-squared tests.

We assessed whether ethnicity associated CpG sites were also cis-expression quantitative trait methylation (cis-eQTMs). *Cis*-eQTM data were extracted from BIOS consortium analyses of methylation and gene expression from blood samples in 2101 Dutch individuals^51^. Enrichment assessed using Wald odds ratio and chi-squared tests.

Biological enrichment analyses were run using unique CpG sites identified from the EWAS and DMR analyses as input. Since hyper- and hypo-methylation at CpG sites in our analysis were likely to be biologically distinct, we stratified our enrichment analyses accordingly. Gene ontology enrichment^81,82^ was conducted using *missMethyl* R package^83^. Further enrichment analyses were conducted using the *LOLA* R package^84^. LOLA input was the genomic coordinates of ethnicity-associated CpG sites and the background set was the genomic coordinates of all array CpG sites included in the EWAS. The LOLA core region set was used to test for enrichments. For each LOLA analysis, results were filtered to retain enriched region sets where the support (i.e. number of regions overlapping) >=5 and the q-value was <0.05.

Analyses were conducted in R, version 4.1.2 (http://www.r-project.org).

## Supporting information

Supplementary Table One

## Data Availability

SABRE data used for this submission will be made available on request to. Further details regarding data sharing can be found on the cohort webpages (https://www.sabrestudy.org/home-2/data-sharing/). Born in Bradford data used for this submission will be made available on request to the Born in Bradford executive committee. The Born in Bradford data management plan (available here: https://borninbradford.nhs.uk/research/how-to-access-data/) describes in detail the policy regarding data sharing, which is through a system of managed open access.

## Authors contributions

HRE conceptualised the study with input from CLR, JLM and KB. HRE led the data analysis and wrote the manuscript. KB conducted genetic PCA analysis. TT, NC and ADH contributed SABRE data. GDS, CLR, NC and ADH contributed to project funding. DAL, DM, GS and JW contributed to the recruitment, data collection and management of BiB data used here. DAL and JW obtained funds for the BiB data used here. All authors contributed to discussion, provided critical input to the project and drafting of the manuscript.

## Acknowledgements

The SABRE study was funded at baseline by the Medical Research Council, Diabetes UK, and the British Heart Foundation. At follow up the study was funded by the Wellcome Trust and the British Heart Foundation. Methylation analysis in the SABRE cohort was supported by a Wellcome Trust Enhancement grant 082464/Z/07/C. Genotyping analysis in the SABRE cohort was supported by the British Heart Foundation (CS/13/1/30327)

Born in Bradford is only possible because of the enthusiasm and commitment of the Children and Parents in BiB. We are grateful to all the participants, teachers, school staff, health professionals and researchers who have made Born in Bradford happen.

BiB receives core infrastructure funding from the Wellcome Trust (WT101597MA), a joint grant from the UK Medical Research Council (MRC) and Economic and Social Science Research Council (ESRC) (MR/N024397/1), the British Heart Foundation (CS/16/4/32482) and the National Institute for Health Research (NIHR) under its Applied Research Collaboration (ARC) for Yorkshire and Humber and the Clinical Research Network (CRN). DNA methylation data was funded by the UK Medical Research Council via the Integrative Epidemiology Unit (MC_UU_12013/5).

HRE, KB, JLM, CLR, DAL and GDS work in the Medical Research Council Integrative Epidemiology Unit at the University of Bristol, which is supported by the Medical Research Council and the University of Bristol (MC_UU_00011/5, MC_UU_00011/6 and MC_UU_00011/1). DAL is supported by a British Heart Foundation Chair (CH/F/20/90003) and National Institute for Health Research Senior Investigator award (NF-0616-10102). NC and AH received support from a Biomedical Research Centre Award to Imperial NHS Healthcare Trust.

The funders played no role in the study design and conduct, or in these analyses or the decision to submit the manuscript for publication. The SABRE study group is entirely independent from the funding bodies.

## List of supplementary Figures

**Supplementary Figure 1.**
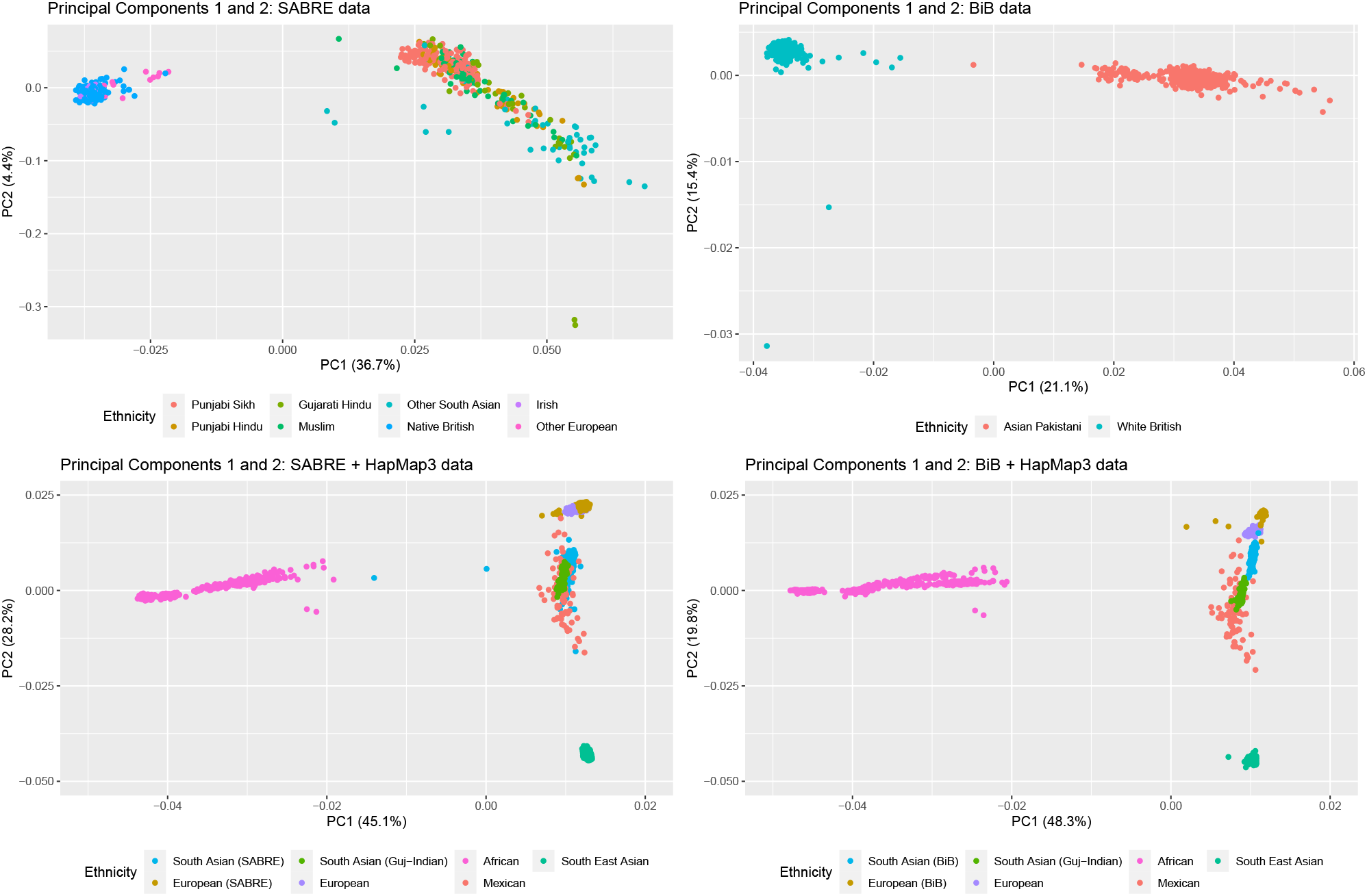
Principal component analysis of SABRE genetic data. Upper panels show PCs 1 and 2 generated from SABRE or BiB data. Lower panels show PCs 1 and 2 generated from SABRE + HapMap3 data or BiB + HapMap3 data. Colours indicate self-reported ethnic group or subgroup (SABRE, BiB) or population group (HapMap3). Axis labels show the variance explained by each PC. HapMap3 populations have been collapsed: South Asian=GIH; European= CEU + TSI; African= ASW + LWK + MKK; Mexican= MEX; South East Asian= CHB + CHD + JPT. In lower panel (SABRE), two outliers self-reporting as South Asian in SABRE data appear intermediate between African groups from HapMap3 and the remaining South Asian cluster. Both of these individuals reported their country of birth as an African country indicating possible genetic admixture in these individuals. These 2 individuals were removed from all other analyses. In lower panel (SABRE), “other South Asian” individuals predominantly identify their country of birth as South Asia (India, Pakistan, Bangladesh, Sri Lanka), n=42/50.

**Supplementary Figure 2.**
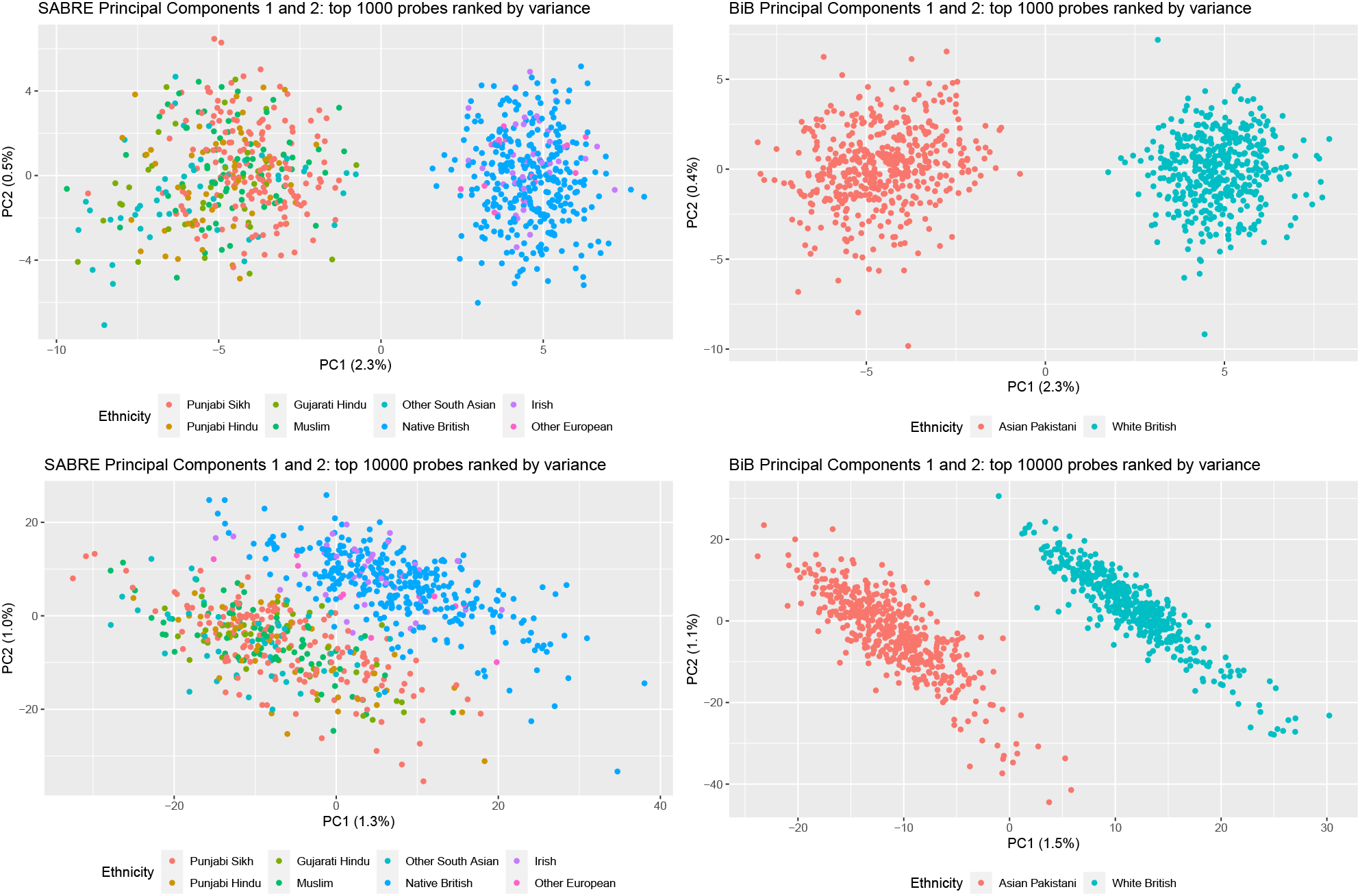
Principal component analysis of SABRE DNA methylation data. Upper panel shows PCs 1 and 2 generated from the 1000 most variant probes in SABRE or BiB. Lower panel shows PCs 1 and 2 generated from the 10000 most variant probes in SABRE or BiB. Colours indicate self-reported ethnic subgrouping (SABRE, BiB). Axis labels show the variance explained by each PC.

**Supplementary Figure 3.**
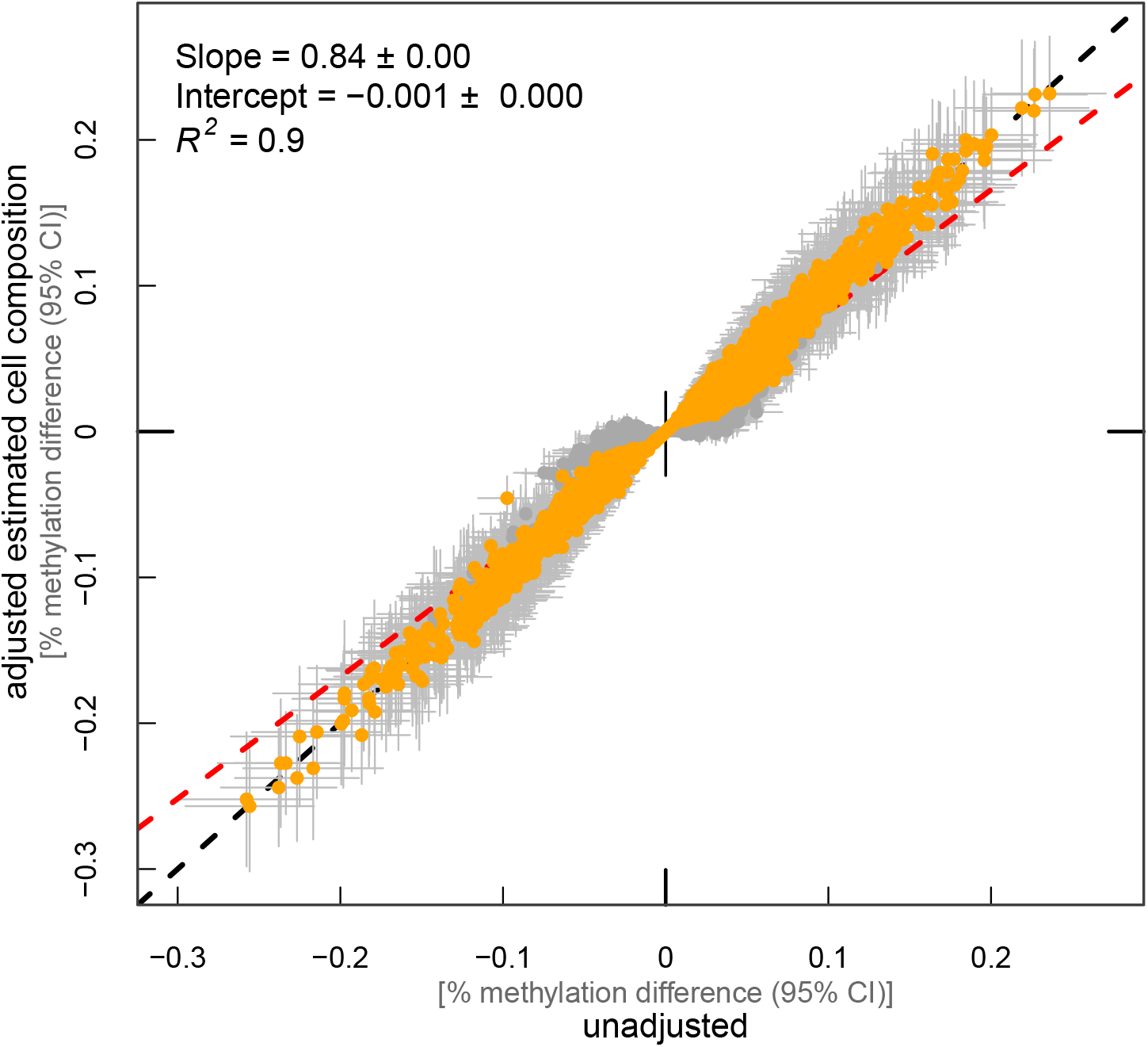
Comparison of effect sizes for cell composition adjustment of EWAS Each CpG is represented by a point on the graph with 95% confidence intervals for effect estimates. Red dashed line: linear regression between datasets. Black dashed line: line of equality. Orange highlighted estimates: p ≤1.03 × 10^−7^ cell adjusted EWAS (n=3922/16,344 CpG sites).

## List of supplementary Tables

Supplementary Table 1 EWAS results

Supplementary Table 2 DMR results

Supplementary Table 3 Epigenetic Age acceleration results

Supplementary Table 4 Cell count differences

Supplementary Table 5 effects of adjustment for mQTL and allele frequency differences

Supplementary Table 6 effects of adjustment for mQTL and allele frequency differences

Supplementary Table 7 ANNOVAR enrichment

Supplementary Table 8 GO term enrichment

Supplementary Table 9 LOLA enrichment

